# Identification of *de novo* mutations in the Chinese ASD cohort via whole-exome sequencing unveils brain regions implicated in autism

**DOI:** 10.1101/2021.07.14.21260545

**Authors:** Bo Yuan, Mengdi Wang, Xinran Wu, Peipei Cheng, Ran Zhang, Ran Zhang, Shunying Yu, Jie Zhang, Yasong Du, Xiaoqun Wang, Zilong Qiu

## Abstract

Autism spectrum disorder (ASD) is a highly heritable neurodevelopmental disorder characterized by deficits in social interactions and repetitive behaviors. Although hundreds of ASD risk genes, implicated in synaptic formation and transcriptional regulation, have been identified through human genetic studies, the East Asian ASD cohorts is still under-represented in the genome-wide genetic studies. Here we performed whole-exome sequencing on 369 ASD trios including probands and unaffected parents of Chinese origin. Using a joint-calling analytical pipeline based on GATK toolkits, we identified numerous *de novo* mutations including 55 high-impact variants and 165 moderate-impact variants, as well as *de novo* copy number variations containing known ASD-related genes. Importantly, combining with single-cell sequencing data from the developing human brain, we found that expression of genes with *de novo* mutations were specifically enriched in pre-, post-central gyrus (PRC, PC) and banks of superior temporal (BST) regions in the human brain. By further analyzing the brain imaging data with ASD and health controls, we found that the gray volume of the right BST in ASD patients significantly decreased comparing to health controls, suggesting the potential structural deficits associated with ASD. Finally, we found that there was decrease in the seed-based functional connectivity (FC) between BST/PC/PRC and sensory areas, insula, as well as frontal lobes in ASD patients. This work indicated that the combinatorial analysis with genome-wide screening, single-cell sequencing and brain imaging data would reveal brain regions contributing to etiology of ASD.

## Introduction

Current prevalence of ASD has approximately increased to 1 in 49 children in the United States, and males are four times more susceptible for ASD than females(1). The epidemiology survey in China showed that the prevalence of ASD may range to 0.2-0.4%, suggesting that the difference in geography and clinical diagnosis criterials may lead to disparate impacts on studies of ASD etiology(2). Recently, tremendous efforts in ASD genetic studies using whole-exome and whole-genome sequencing have built up high-throughput assessment pipelines for protein-disrupting variants in large ASD cohorts, in which *de novo* single nucleotide variants (SNVs), insertions and deletions (INDELs) and copy number variants (CNVs), as well as rare inherited variants are major contributors for genetic risks of ASD(3-6). Thus it is critical to further classify genetic causes from accumulated ASD genetic studies in consideration of neurobiological evidences. The online SFARI gene database provided an important public resource for ASD risk genes, in which over one thousand of ASD candidate genes were prioritized with genetic and neurobiological evidences (Category S, 1, 2, 3) (7).

Although genomic information of large cohorts consisting of tens of thousands ASD patients have been collected, East Asian populations are still underrepresented groups(8). Some genetic studies on Chinese ASD cohorts using targeted multiplex sequencing technology focused on a group of genes associated with neurodevelopmental disorders, which cannot yield comprehensive genome-wide information about ASD risk genes(9, 10). Two available genetic studies on Chinese ASD cohorts using whole-genome sequencing methods included less than 40 trios, limiting the power of genomic sequencing(11, 12).

In this study, we performed whole-exome sequencing analysis in a Chinese cohort including 369 ASD probands with their parents. 150 bp paired-end sequencing short reads were mapped against human reference genome build 38 (GRCh38/hg38). SNVs and INDELs were jointly called across all samples and filtered by GATK Variant Quality Score Recalibration (VQSR) and Convolutional Neural Network (CNN) tools. Together with analysis of single-cell sequencing data from the developing human brain, we found that expression of genes with *de novo* mutations were specifically enriched in pre-, post-center gyrus (PRC, PC) and BST (banks of superior temporal) regions in the human brain.

By further analysis of the brain imaging data with ASD and health controls, we found that BST of the right hemisphere in ASD patients significantly decreased gray volume comparing to health controls, suggesting the potential structural deficits associated with BST in autistic patients. Finally, after analyzing the seed-based functional connectivity (FC) of these regions, we found the decrease in FC between BST/PC/PRC and sensory areas, insula, as well as frontal lobes in ASD. This work indicated that the in-depth combinatorial analysis of ASD risk genes from genome-wide screening, the single-cell sequencing and brain imaging data would unveil the brain regions implicated in ASD and thus provide an analytical framework illustrating the genetic basis and neurobiological mechanism for ASD.

## Results

### Identification of *de novo* variant in ASD probands

We analyzed a ASD cohort consisting of 369 ASD probands and 706 parents from 353 pedigrees recruited from Department of the Child and Adolescent Psychiatry, Shanghai Mental Health Center. Among the cohort, there are 15 multiplex family containing two ASD children and 338 simplex family which have one ASD child. The fourth edition of the Diagnostic and Statistical Manual of Mental Disorders (DSM-IV) were used for ASD diagnoses by trained psychiatrists.

Proportion of the targeted exome regions covered with ≥ 20x or 40x of reads indicates sufficient coverage (Fig. S1A). After performing the multidimensional scaling of the genotyping data of common exonic SNPs was performed by using PLINK (a whole genome association analysis toolset)(13), we found that all probands in this cohort were included in the cluster of East Asian individuals (Fig. 1A).

**Figure 1.**
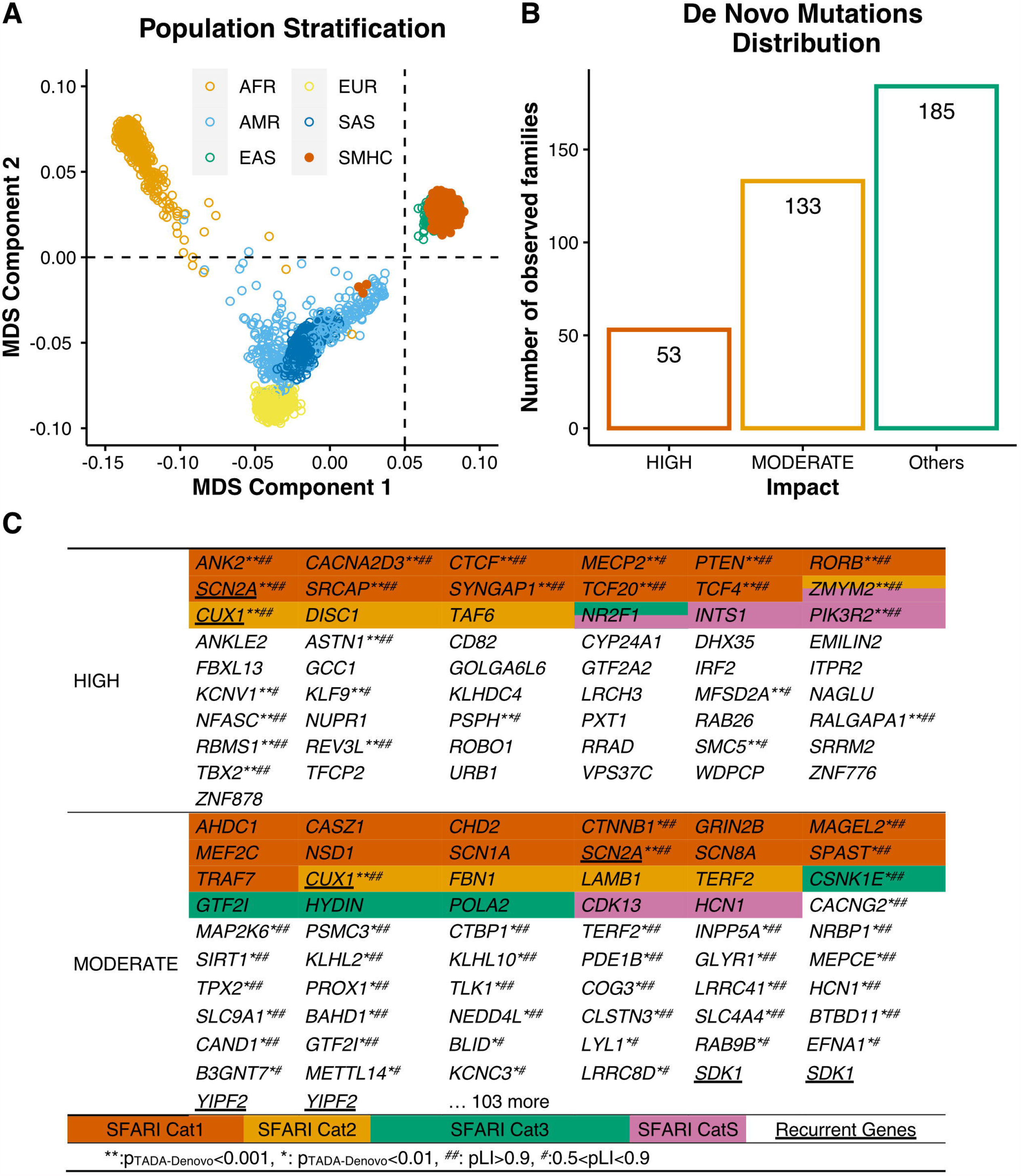
Identification of *De Novo* Mutations in ASD Probands. (A) Multidimensional scaling plots of 369 ASD probands in our cohort (OWN) along with African (AFR), American (AMR), East Asian (EAS), European (EUR) and South Asian (SAS) individuals in the 1000 Genomes Project. Figure was generated by analyzing genotyping data of 1,064 common exonic SNPs using PLINK. The first and second dimensions are shown. The red, blue, green, magenta and yellow circles dots indicate AFR, AMR, EAS, EUR SAS individuals. The orange dots indicate ASD probands exome sequenced in this study. (B) The number of families with various types of *de novo* mutations in our ASD cohort. Others stand for no detection of High- or Moderate-impact *de novo* mutations. (C) The list of High-impact and Moderate-impact mutations identified in this ASD cohort. The gene also presented in the SFARI gene list were highlighted with various colors representing 4 catogeries (Cat S, 1-3).

After performing variant filtering, we discovered a set of 442 *de novo* mutations (DNMs) (Table S1). We classified DNMs into three categories, including High-impact, Moderate-impact, and Possible-damaging. The High- and Moderate-impact were defined by VEP (Ensembl Variant Effect Predictor, https://asia.ensembl.org/info/docs/tools/vep/index.html). Briefly, the High-impact variants usually lead to truncation of protein products, including gain or loss of STOP codons as well as frameshift-causing insertions and deletions (INDELs). Interestingly, among the 55 genes containing High-impact SNVs, there are 18 genes previously reported in the SFARI gene list (Category S, 1, 2, 3) such as *SCN2A, PTEN, MECP2, SRCAP, TCF4*, suggesting that there are substantially non-SFARI ASD gene in the Chinese cohorts (Fig. 1B, C).

Moderate-impact variants were defined as protein sequence changing, but not truncating, such as missense SNVs and inframe INDELs. To further categorize the severity of missense variants, we annotated missense SNVs into a new class, named Possible damaging missense DNMs, which were defined as the variants predicted to be damaging by at least two of the seven following prediction algorithms: SIFT(14), PolyPhen-2 HumVar(15), PolyPhen-2 HumDiv(15), LRT(16), Mutation Taster(17), Mutation Assessor(18) and PROVEAN(19) annotated by dbNSFP4.0a(20, 21). Interestingly, among 165 Moderate-impact variants, there are only 23 variants are present in the SFARI gene list (Fig.1 B, C).

Over one thousand ASD risk genes in the SFARI gene list were mainly found from genetic studies in US and European studies, therefore we were wondering whether numerous genes with DNMs in the Chinese ASD cohorts which were not included in the SFARI list were really contributory to ASD or some common genetic variants may not associated with disorders. To further determine whether these genes with DNMs may be contributory to ASD, next we statistically evaluated the contributions of each *de novo* variant to ASD using the Transmission and *De Novo* Association Test-Denovo (TADA-Denovo) method. We first measure the frequence of *de novo* and missense variants in each gene with DNMR-SC-subtype data(22), then applied the TADA-Denovo method(23). We classified the DNM variants with p values obtained from TADA-Denovo test into two tiers (*, *p* < 0.01, or **, *p* < 0.001) (Table S2). We further measured the “probability of loss-of-function intolerance” (pLi) score for each variant and categoried variants with signifant TADA-Denovo value into two tiers as well (>0.9 represented by ^##^, 0.5-0.9 represented by ^#^)(24). Finally, we found that 11 genes with High-impact mutations and 35 genes with Moderate-impact mutations, all of them not included in the SFARI gene list, were signficanlty with both TADA-Denovo and pLi score, further strengthen their contributions to ASD (Fig. 1C).

We would like to investigate whether genes with *de novo* variants identified in various ASD genetic studies may be overlapping. Interestingly, we found that *de novo* ASD risk genes detected in ASD probands in this sduty (the SMHC cohort) showed little overlapped with the list of *de novo* ASD risk genes from the Japanese cohort(Fig. S1B, C)(8). Moreover, we found that there were also little overlapping in *de novo* variants between the SMHC cohorts with other studies with 200-400 trios (Fig. S1D)(5, 25-27).

### Identification of *de novo* CNVs in ASD risk genes with the WES dataset

Although the gold standard for copy number variations detection is the chromosomal microarray analysis (CMA), various toolkits has emerged to identify CNVs with the whole-exome sequencing (WES) dataset(28). However, the current reported algorithms for CNV detection is not optimal for the WES dataset and incompatible with the GRCh38/hg38 reference genome.

We applied a germline CNV calling protocol based on GATK cohort mode (version 4.2.0.0) (See Supplementary Methods) and identified numerous *de novo* CNVs in the probands (Fig. 2A-N, Table S3). To exclude the false positive hits, we set 2 standards for CNV screening. First, selection of duplication or deletion signals appearing in more than 2 continuous exons. Second, CNVs should fulfill the HIGH-impact criterial, leading to protein truncation, such as deletion of START or STOP codons.

**Figure 2.**
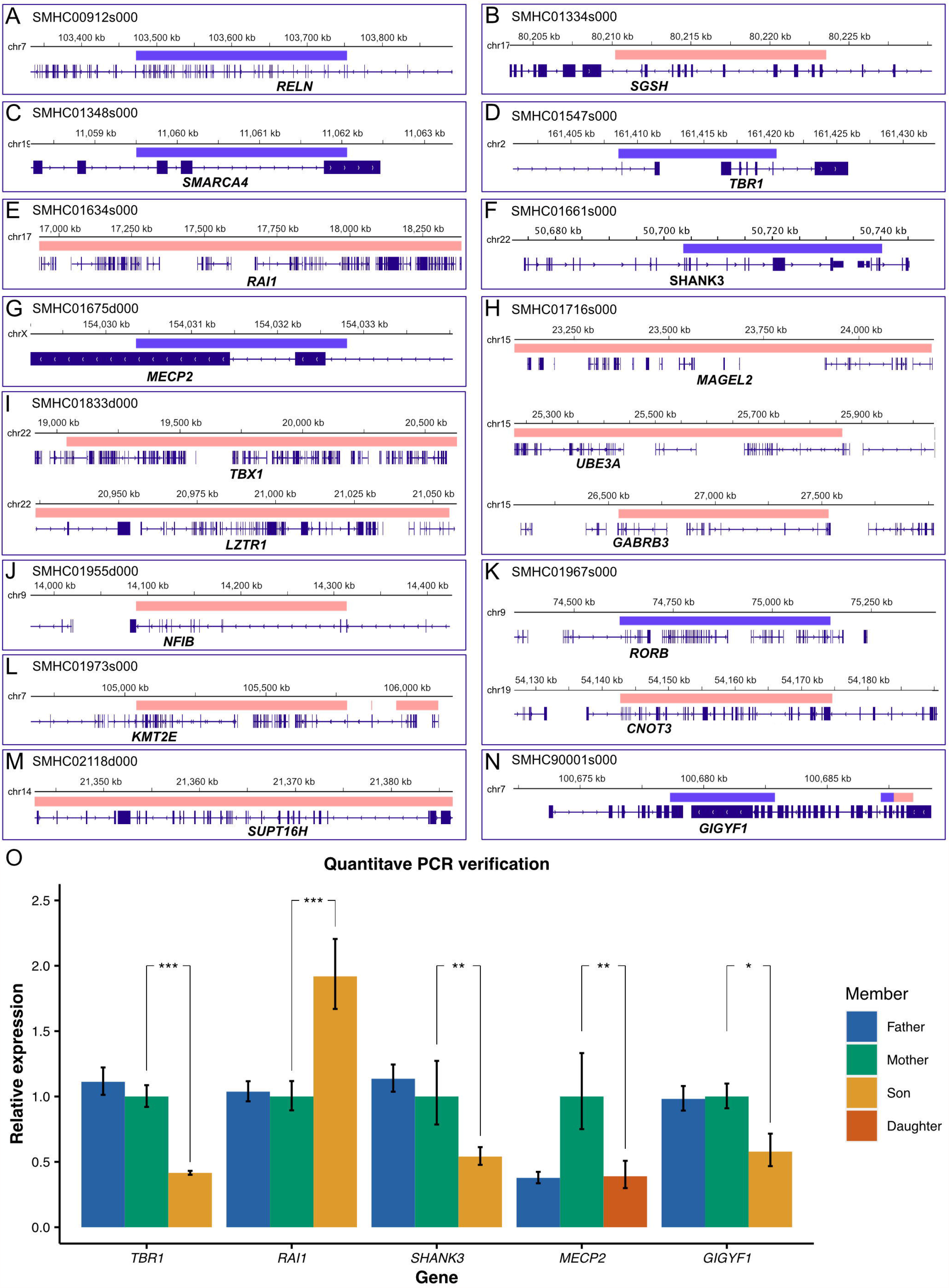
Identification of *De Novo* CNVs. (A-N) Schematic diagrams of 18 SFARI ASD risk genes (Cat S: 4 genes, Cat 1:14 genes) with *de novo* CNVs. Blue: deleted chromosomal segment. Red: duplicated chromosomal segments. (O) Quantitative real-time PCR verification of *de novo* CNVs. *TBR1*, t-test, p=3.22×10^−7^; *RAI1*, t-test, p=7.40×10^−7^; *MECP2*, t-test, p=0.0069; *SHANK3*, t-test, p=0.0028. All experiments were repeated more than 4 independent times. Error bars represent standard errors from four or six replicates.

To prioritize ASD risk genes, we first examine CNVs happened in the known SFARI genes (Fig. 2A-N). We found 18 CNVs exhibiting duplication or deletions in known SFARI genes (Cat S:4 genes, Cat 1:14 genes), such as duplications of *RAI1, UBE3A* and deletions of *TBR1, SHANK3, MECP2, GIGYF1* (Fig. 2A-N). We further validated the CNV results by performing quantitative PCR, confirming the feasibility and faithfulness of our new methods (Fig. 2O).

Furthermore, among *de novov* large CNVs we found, there are 9 CNVs containing genes in the SFARI Cat 2 gene list (Table S3). There are totally 26 CNVs containing critical ASD-risk genes in the SFARI gene list (Cat S, 1, 2), suggesting that genes implicated in these *de novo* large CNVs may contribute to pathogenesis of ASD.

### Expression of ASD risk genes enriched at PC, PRC and BST regions in the developing human brain

The etiology of ASD may be disruption of neural circuits associated with social behaviors, thus identification of the expression profile of gene with DNMs in the human brain would provide critical insights for which brain regions may be affected by mutations of ASD risk genes(29). To acquire the expression pattern of ASD risk genes in the single-cell resolution, we used the recent single-cell sequencing database in the developing human brain(30, 31). We grouped total 17434 transcriptomes collected from gestational week (GW) 09-26 of human fetus brains and catogeried them into sub-cell types according to marker genes (Fig. 3A, Fig. S2A-C).

**Figure 3.**
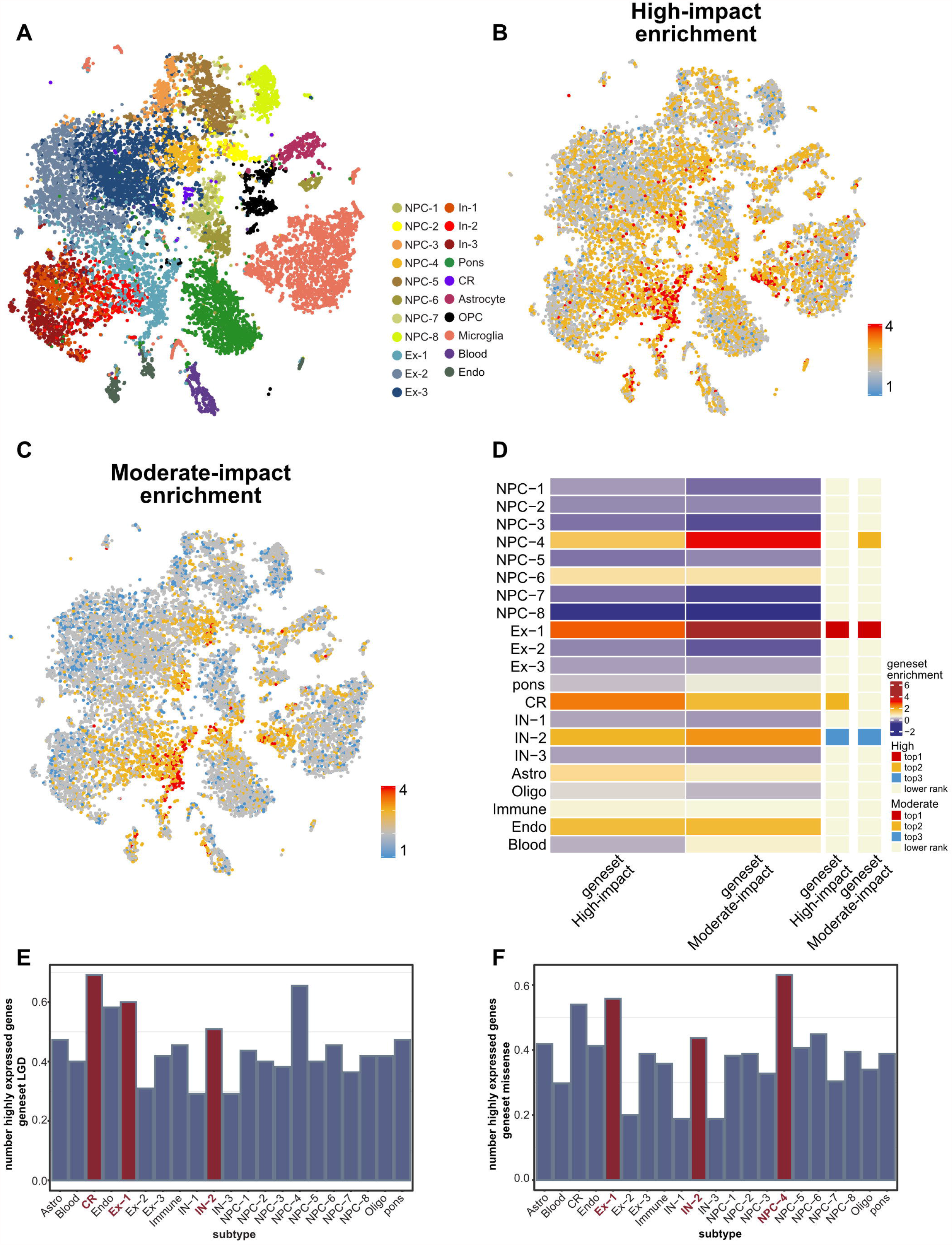
Expression pattern of ASD risk genes among different cell types. (A) clustering of single-cell RNA-seq data from different brain regions. Cell types were colored differently. (B) Visualization of enrichment score of the 55 High-impact genes by UMAP. (C) Visualization of enrichment score of the 165 Moderate-impact genes by UMAP. (D) Heatmap showing averaged enrichment score of ASD risk genes among different subtypes (high, red; low, blue). The top three most enriched cell types for each gene-set were shown on the sidebar. (E) Histogram showing the proportion of highly expressed genes (genes expressed in at least 25% cells from individual cell type) of the 55 High-impact genes among cell types. (F) Histogram showing the proportion of highly expressed genes (genes expressed in at least 25% cells from individual cell type) of the 165 Moderate-impact genes among cell types.

We first examined the expression pattern of 55 High-impact genes and 165 Moderate-impact genes in various cell types, and found that both High-impact genes and Moderate-impact genes were highly expressed in several subtypes of cells, including NPC-4, Ex-1 and In-2, as well as cajal-retzius cells (CR) (Fig. 3B, C, D). We further looked into where NPC-4, Ex-1, In-2, and cajal-retzius cells (CR) localized in the developing human brain. We found that NPC-4 was generally distributed in the four major lobes of the brian, suggesting that this specific sub-group of neural progenital cells may be associated with ASD (Fig. 3E, F, Fig.S3A, B). However, Ex-1 and In-2 specifially enriched in some sub-regions of the brain including precentral gyrus (PRC), postcentral gyrus (PC) and banks of superior temporal sulcus (BST) regions (Fig. S3C-E).

We next investigated whether expressions of ASD risk genes may be enriched in specific brain regions of the human brain. In previous work, the single-cell sequencing were preformed in 22 brain subregions in the developing human brain (Fig. 4A)(30). Surprisingly, we found that the High- and Moderate-impact genes were significantly enriched in precentral gyrus (PRC), postcentral gyrus (PC) and banks of superior temporal sulcus (BST) regions (Fig. 4B, C, D, E). The PRC is the primary motor cortex (M1) and PC is the primary somatosensory cortex (S1). The implications of PRC and PC in ASD had been reported previously(32, 33). Interestingly, we also found the functional connectivity including right S1 (S1R) and M1 (M1R) regions were specifically decreased in *MECP2* transgenic monkeys, the non-human primate model for autism, comparing to wild-type monkeys (34-36).

**Figure 4.**
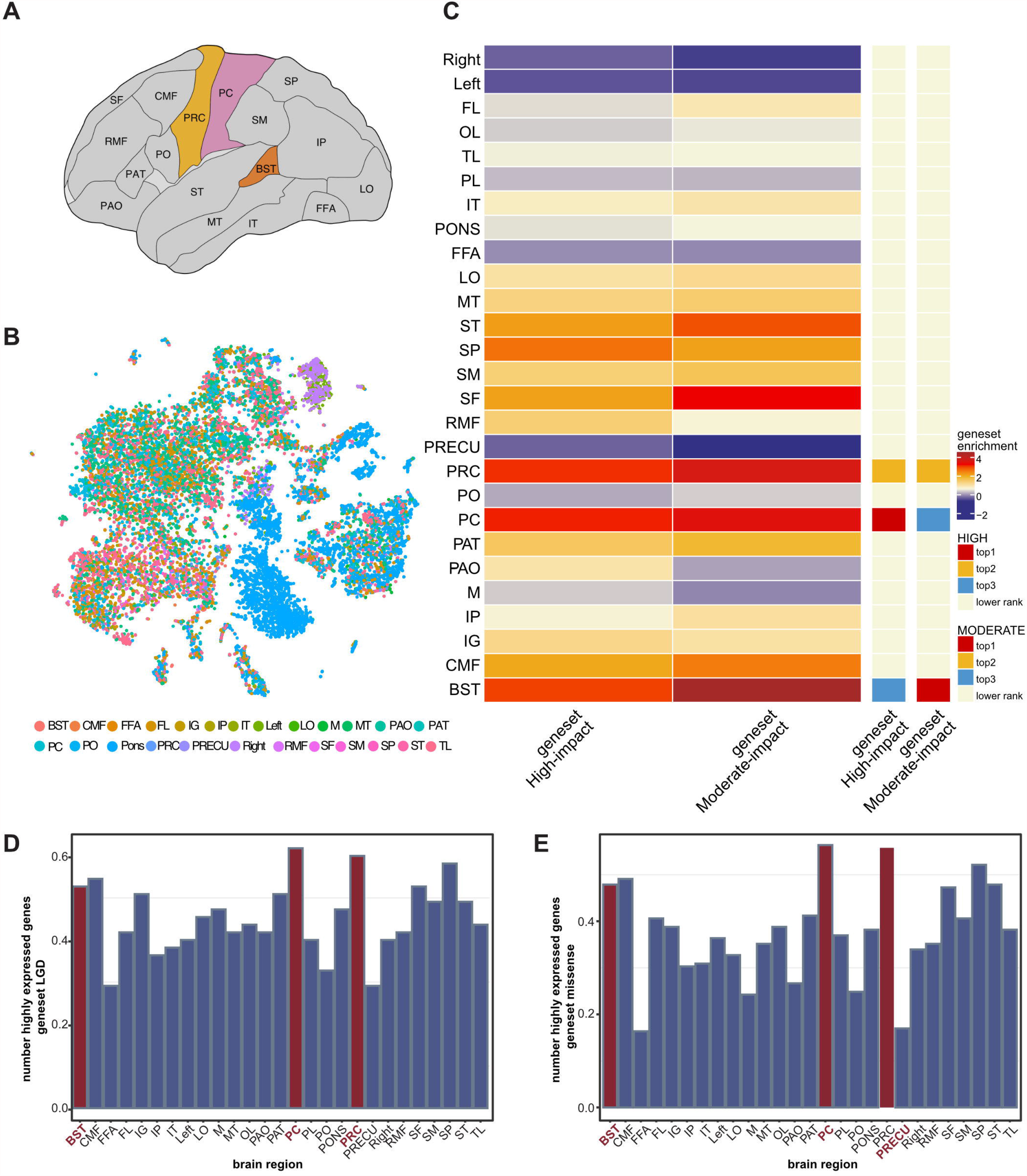
Expression pattern of ASD risk genes among different brain regions. (A) The schematic diagram showing the 22 brain sub-regions used in published human single cell RNA-seq data was applied in analysis of this work. (B) UMAP displaying diverse brain regions (Abbreviations are listed in Table S7). Brain sub-regions were differently colored. (C) Heatmap showing averaged enrichment score of ASD risk genes among different brain regions (high, red; low, blue). The top three most enriched brain regions for each gene-set were shown on the sidebar. (D-E) Histogram displaying the proportion of highly expressed genes (genes expressed in at least 25% cells from individual brain region) of the 55 High-impact genes (D) and the 165 Moderate-impact genes (E) among various brain sub-regions.

### Brain imaging analysis

In order to determine whether these brain regions were affected in ASD patients from differerent populations, we acquired imaging data from Autism Brain Imaging Data Exchange (ABIDE-I, http://fcon_1000.projects.nitrc.org/indi/abide/)(37) a publicly available database released containing 1112 subjects (539 ASDs, 573 age-matched healthy controls-HCs) from 16 international imaging sites underwent anatomical and resting-state functional MRI scans. we collected more than 200 age-matched brain imaging data from ASD or HC groups of ABIDE-I (Table S4).

To further validate whether PC/PRC and BST may have structural (gray matter) alternations in ASD patients, we first performed the voxel-based morphometry (VBM) analysis of these region in ASD and HC using T1 data. Surprisingly, we found that the gray matter volume of BST in the right hemisphere was significantly smaller in the ASD group than that in the HC group (t = 3.61, p = 0.003, t- and p-value from linear mixed model detailed in Statistic section of Methods and Materials), and this effect persisted even after controlling for medication status (t = 3.32, p = 0.001) and full-scale intelligence quotient (FIQ) (t = 3.4, p = 0.0007) (Fig. 5A, B, C).

**Figure 5.**
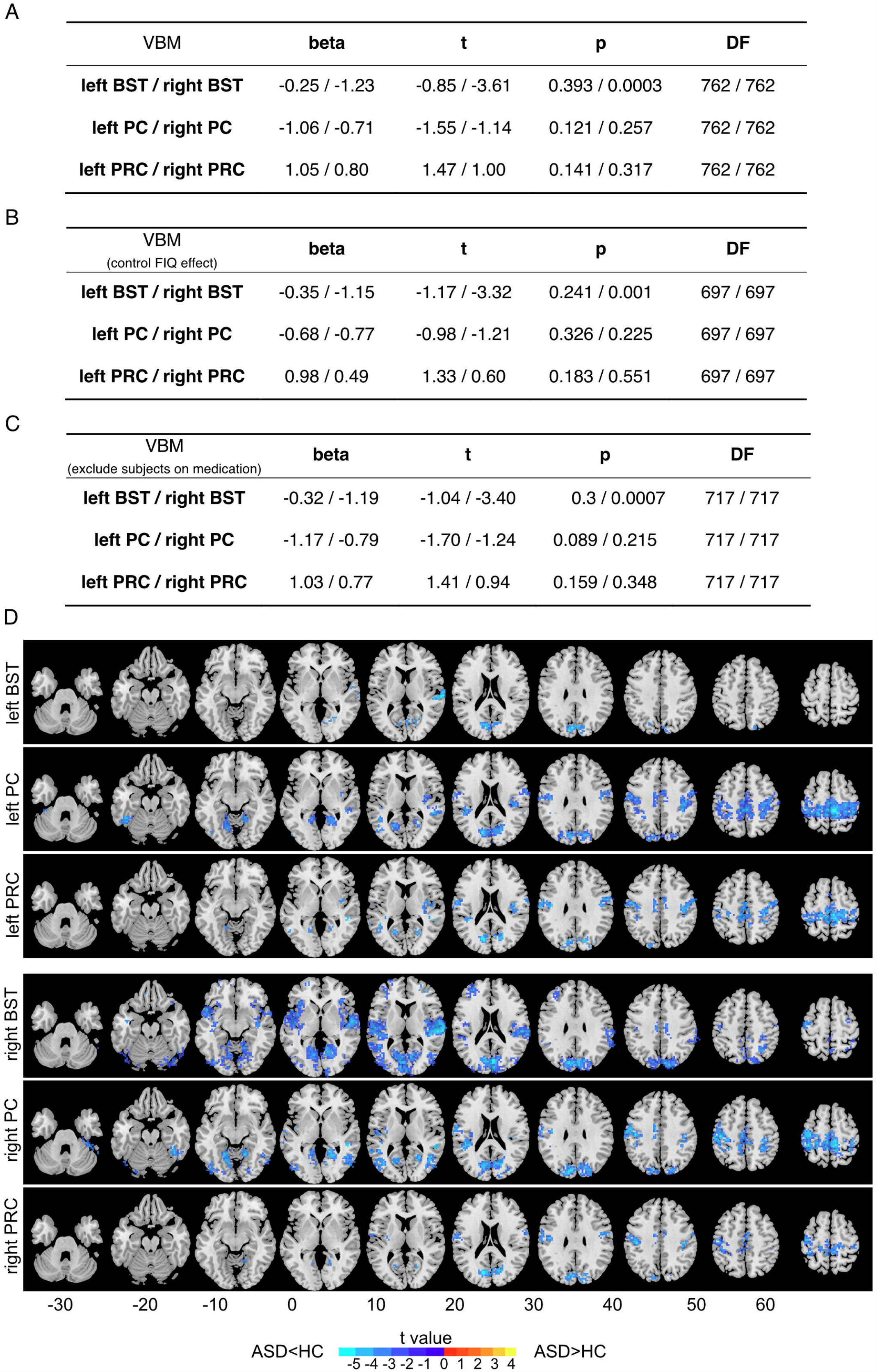
Structural and functional connectivity between ASD and healthy controls in PC, PRC and BST regions. (A) VBM. Beta, t, p, and degree of freedom (DF) are obtained from linear mixed models. Beta is the regression coefficient of the fixed effect. DF is the degree of freedom of the model (equal to the sample size minus the number of parameters to be estimated). T stands for t-statistics for testing the null hypothesis that the coefficient is equal to zero. P stands for p-value for the t-test. (B) VBM data with control of FIQ effect. (C) VMB data excludes subjects with medications. (D) Group differences between ASD and healthy controls in seed-based resting-state Functional Connectivity(FC). 6 brain regions including bilateral BST, PC and PRC were selected as seeds. Functional connectivity between these 6 regions and the whole-brain voxels were calculated. Only the clusters of voxels with significant FC differences between the two groups (the two-sample t-test, pFWE < 0.05, cluster size < 20 voxels or 180 mm^3^) were displayed.

We finally investigate the potential functional connectivity (FC) between the above regions of interests (ROIs) and the whole-brain voxels, by performing seed-based FC analysis using resting-state functional MRI scans data from ASD and HCs. Consistently, we observed a significant decrease in connectivity between BST/PC/PRC and sensory areas, insula, as well as frontal lobes in ASD compared to HC (Fig. 5D). We found a decrease of all six ROIs’ functional connectivity to the occipital lobe region, which is commonly associated with vision. We also found decreased connectivity between bilateral PC/PRC to the sensorimotor region of the parietal lobe. In addition, on the right BST, we found the most widely FC decrease among all ROIs, including connections to the right insula and temporal lobes (t = −6.05, FWE corrected p = 0.0002), to the bilateral frontal lobe and to the occipital lobe (Table S5-S6 and Fig. 5D).

BST has been shown to be voice-selective areas in normal adults, which plays a role in voice recognition and social stimuli processing (38). In an fMRI study, activation of BST by speech stimulation appeared compromised in adults with ASD (39). In addition, BST also exhibited ASD-related functional connectivity alterations (40, 41), gray matter changes (such as lower surface area and greater age-related cortical thinning) (42, 43) and white matter volume reduction (44). Our study indicated that genetic predepositions in ASD patients may lead to structural and function abnormalities in brain regions associated with processing of social information, thus providing novel candidate brain regions for intervention of ASD.

## Discussion

With accumulating genomic studies on autism cohorts world-wide, the genetic architecture of ASD has emerged over the last decade. Composed of *de novo* and rare inherited mutations, genetic variants play a decisive role in determining the etiology of ASD. Although the rapid development of DNA sequencing technology, precise identification of genetic variants in the large scale genome sequencing over hundreds and thousands of ASD core trios is still very challenging.

In this work, we applied the latest GATK package (v4.1.4.1) and the GRCh38/hg38 dataset, which is compatible for ongoing update of Ensembl genome database. We focused on the identification of *de novo* variants, including SNVs, INDELs and CNVs, with the customized joint calling pipeline. Importantly, we found several critical CNVs containing ASD-risk genes, such as *SHANK3, TBR1* and *MECP2*, indicating that screening CNVs with the WES dataset would be very valuable for ASD genetic studies. Interestingly, about 30% (18/55) genes carried *de novo* High-impact variants existed in the SFARI gene list, suggesting that there are potentially novel ASD genes in the Chinese cohorts. With the in-depth analysis of TADA-Denovo and pLi evaluation, we found that there are a substantially portion of DNM discovered in the Chinese ASD cohorts appeared to be significant statiscally. Taken together, we suggest that although the overall genetic architecture of ASD remains similar across different populations, the frequence of individual genetic component may vary due to geographic isolations. Thus, in order to comprehensively acquire the ASD risk genes, genome-wide sequencing in large cohorts from different populations would be required.

One of intriguing hypothesis of etiology of ASD is that genes carrying genetic mutations in ASD patients may express in some specific brain regions governing social behavior-related circuits. Thus precise idenfication the expression pattern of ASD risk genes in human brain, with single-cell resolution, would be an ideal approach. In this work, we took advantage of single-cell sequencing database collected from various brain regions across gestational week 9-26 and found that the expression profiles of ASD risk genes in the developing human brain indeed exhibited specific patterns. ASD risk genes discovered in the Chinsese ASD cohort specifically enriched in the primary somatosensory (S1-PC) and primary motor cortices (M1-PRC), as well as the BST region.

Although there are evidences suggesting that S1 and M1 cortcies showed defects in functional connectivity in ASD samples (34-36), it still difficult to determine whether the defects associated with S1 or M1 are the cause or conseqeunce of deficits in social behaviors. It is well known that ASD patients often exhibited abnormal somatosensory functions. Thus our data suggested that abnormalities in somatosensory functions may root from central control, other than peripheral functions.

The finding that right BST region is implicated in ASD pathogenesis is extremely intriguing. The brain regions in the right hemisphere are stronger associated with autism than left hemisphere are also obversed in previous work (34-36). The connections between right-hemisphere with autism are also discussed(45). Involvement of BST in social perception has been found in previous work with human subjects(46). Thus we hypothesized that dysregulation of ASD risk genes in BST likely caused abnormalities in social perception and related functions. Since BST is on the surface region of human brain, one may be able to design neuromodulation methods to activate the neural activity in the right BST region of ASD patients, through transcranial electrical or magnetic stimulations.

Taken together, this work presented numerous ASD risk genes from whole-exome sequencing of 369 Chinese ASD cohorts. Beside known ASD candidate genes present in the SFARI gene database, we further measured the probability of DNMs contributions to ASD through the TADA-Denovo test and found that quite a few ASD candidate genes appeared to be statiscally significant, suggesting that whole-exome sequencing on large ASD cohorts are indeed valuable to elucidate the genetic landscape of ASD in different populations. Importantly, the combinational analysis of single-cell sequencing and brain imaging in this work presented an analytical framework in which one could address the potential etiology of ASD from genetic discoveries.

## Data Availability

The datasets used and/or analysed during the current study are available from the lead contact on reasonable request.

## List of abbreviations

ASD: Autism Spectrum Disorder
SFARI: Simons Foundation Autism Research Initiative
SNVs: single nucleotide variants
INDELs: insertions and deletions

The abbreviations for brain subregions are listed in Table S7.

## Ethics approval and consent to participate

Experiments were approved by the Institutional Review Board (IRB), Shanghai Mental Health Center of Shanghai Jiao Tong University (FWA number 00003065; IROG number 0002202). Ethical review number of this study is 2016–4, and committee members of IRB who approved this study was Dr. Yi-Feng Xu. Patients were collected from outpatient Department of the Child and Adolescent Psychiatry, Shanghai Mental Health Center. Written informed consent forms were obtained from parents for all minor children. All sample collections were handled according to the protocol approved by the Institutional Review Boards.

## Acknowledgements

The authors thank the families for their participation in this study. This work was supported by grants from the NSFC Grants (#31625013, #81941405, #32000726); Shanghai Brain-Intelligence Project from STCSM (16JC1420501); Strategic Priority Research Program of the Chinese Academy of Sciences (XDBS01060200); Program of Shanghai Academic Research Leader, the Open Large Infrastructure Research of Chinese Academy of Sciences, and the Shanghai Municipal Science and Technology Major Project (#2018SHZDZX05). J.Z. was supported by NSFC 61973086, and Shanghai Municipal Science and Technology Major Project (No.2018SHZDZX01).

## Authors’ contributions

All authors contributed to the work and meet the criteria for authorship. Study design: B Yuan and Z Qiu. Experiment and data analysis: B Yuan, M Wang, X Wu. Acquisition of Clinical information: PP Cheng, R Zhang (SMHC), Y Yu, YS Du. Interpretation of WES data: B Yuan. CNV validation experiment: B Yuan, PP Cheng, R Zhang (ION). Analysis of single-cell sequencing data: M Wang, XQ Wang. Analysis of brain imaging data, X Wu, J Zhang. Drafting of the manuscript: B Yuan, Z Qiu. Study supervision: Z Qiu, YS Du, XQ Wang, J Zhang.

---

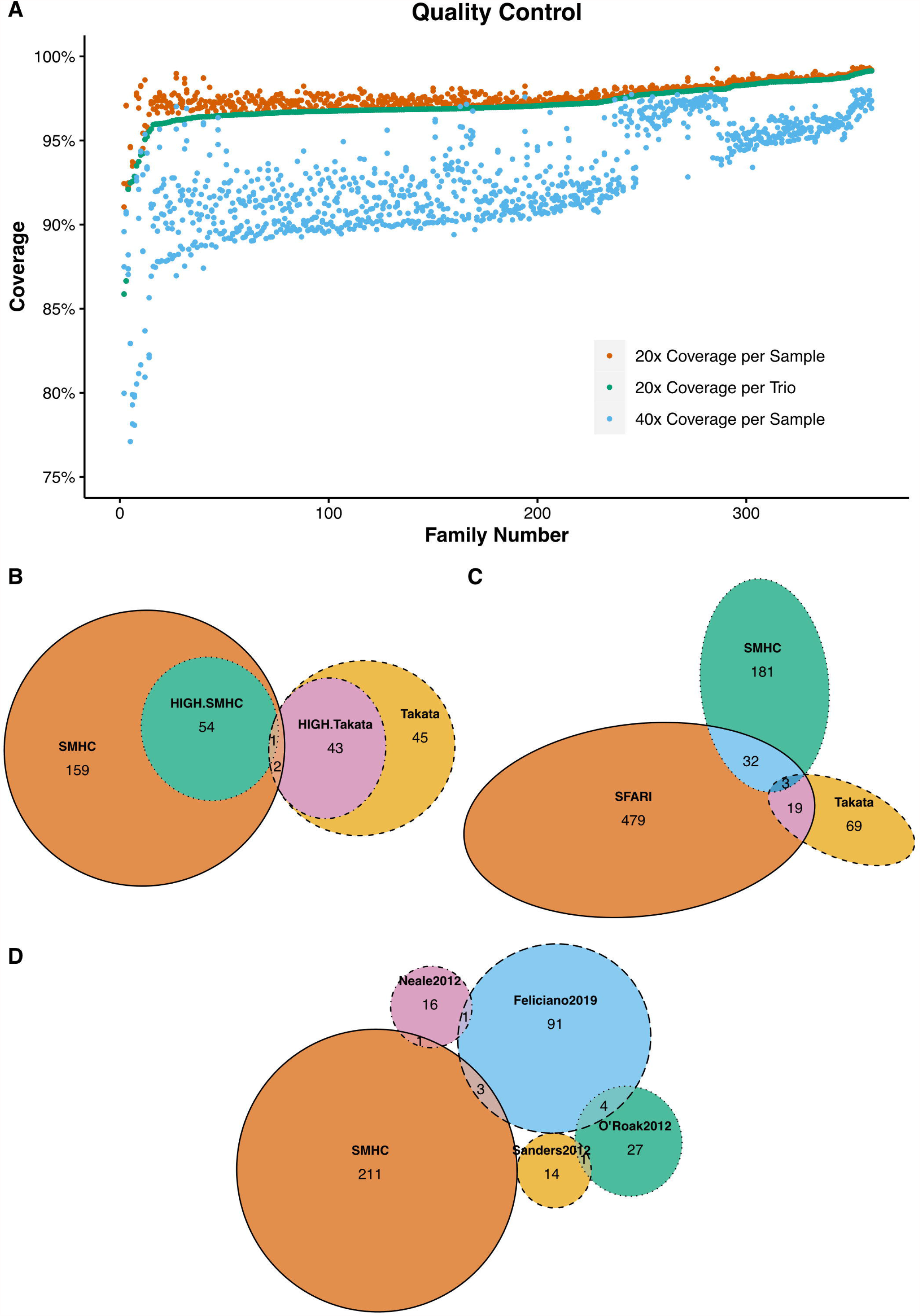

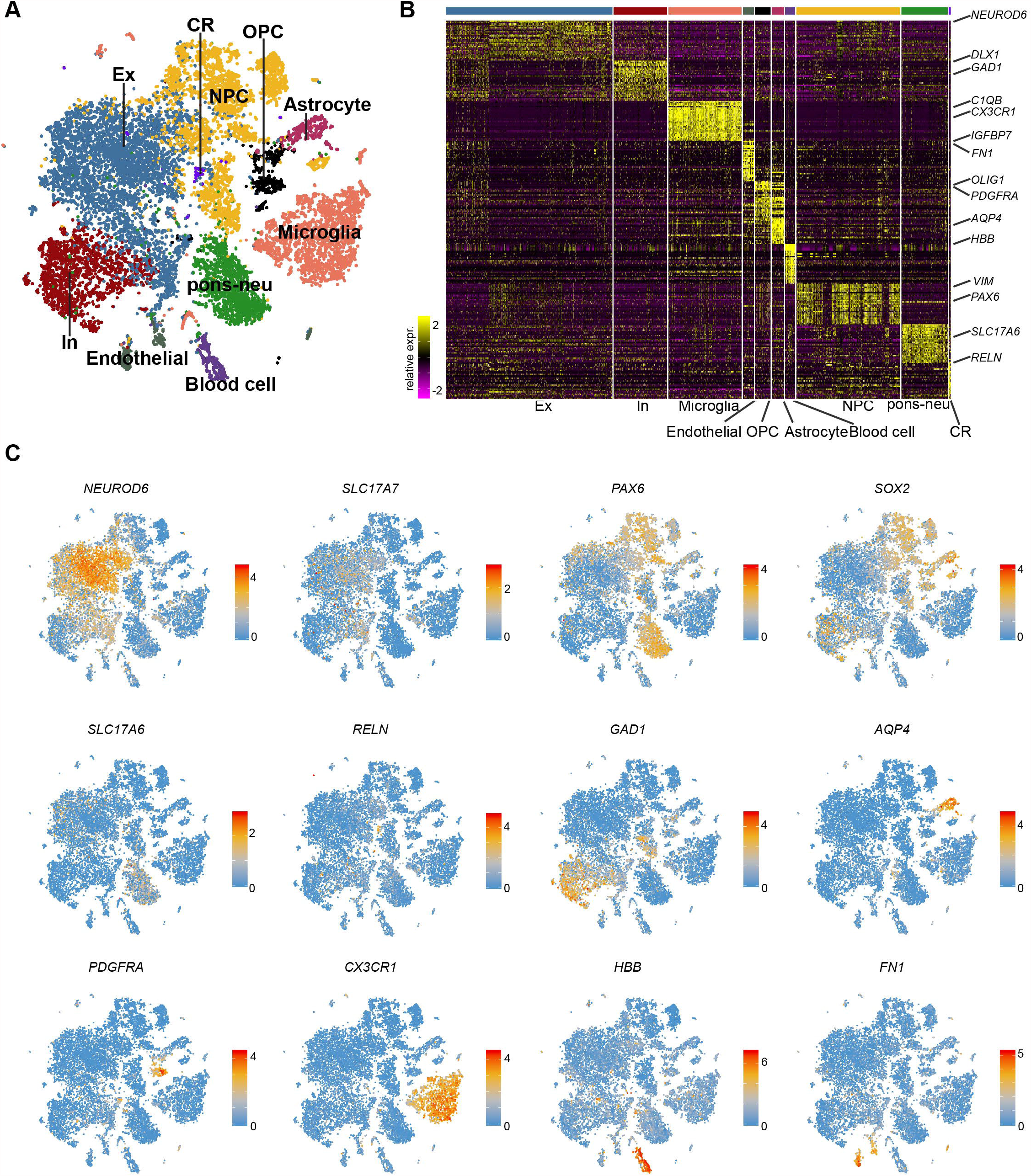

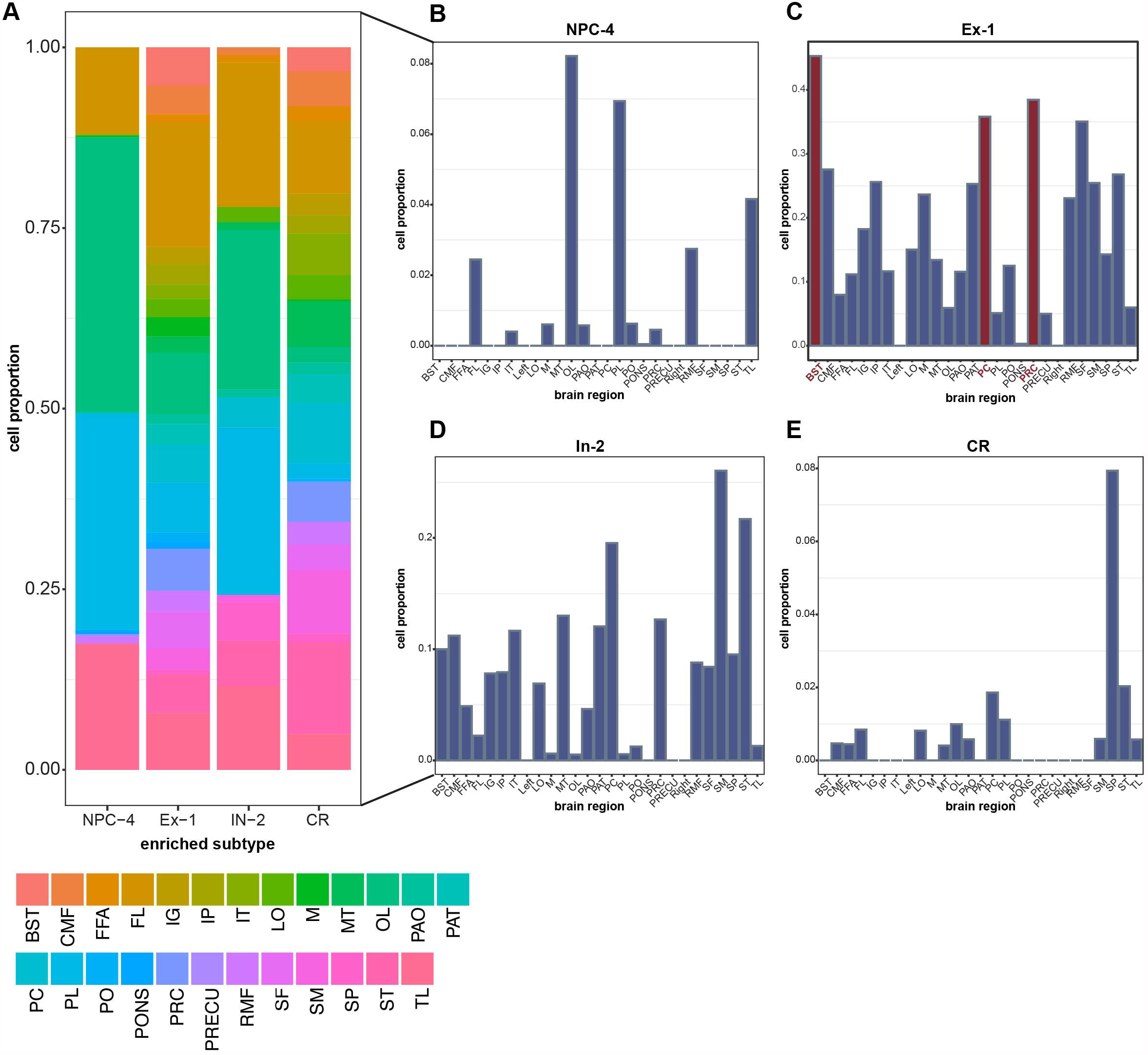

