## Supplemental Materials and Methods for "Identification of *de novo* mutations in the Chinese ASD cohort via whole-exome sequencing unveils brain regions implicated in autism"

### Supplemental Figure 1-3 legends

**Table S1-S7**

### Supplemental Materials and methods

**Supplemental Figure legends**

**Figure S1. Quality Control Results and Enrichment Analyses of Various Functional Types of *de novo* mutations in Our and Published ASD Cohorts**

(A)Sequencing coverage performance. Proportion of the target exome region covered with the indicated numbers (≥ 10x or 30x) of reads was plotted. Data were sorted in the order of the 10x trio rank (trios with the largest proportion covered with ≥ 10x reads on the left and the smallest on the right). Red dots indicate 10x individual coverage and blue dots indicate 30x individual coverage.

(B) The overlap of ASD risk genes between the Chinese ASD cohort (SMHC) and Japanese ASD cohort (Takata). The dark yellow circle indicates genes with DNMs in the Chinese ASD cohort; the light yellow circle indicates genes with DNM mutations confirmed by Sanger sequencing in the Japanese ASD cohort; the green and magenta circles indicate genes with HIGH-impact DNMs in Chinese and Japanese ASD cohorts, respectively.

(C) The overlap of ASD risk genes between the Chinese ASD cohort (SMHC), the Japanese ASD cohort (Takata) and the SFARI gene list (including Cat S, 1, 2). The green and yellow circles indicate genes with DNMs in Chinese and Japanese ASD cohorts.

(D) The overlap of ASD risk genes between the Chinese ASD cohort (SMHC) with four other studies (samples size from 200-400 ASD trios). All genes with DNM were analyzed.

**Figure S2 Overview of transcriptional heterogeneity among cells from diverse brain regions.**

(A) Visualization of major cell type by UMAP. (B) Heatmap illustrating the differently expressed genes among major cell types. Well-known marker genes for each cell type were labeled on the left side of the heatmap (high, yellow; low, purple). (C) UAMP showing the expression of marker genes. *NEUROD6* and *SLC17A7* for excitatory neurons; *PAX6* and *SOX2* for NPCs; *SLC17A6* for neurons from pons; *RELN* for Cajal-Retzius cells; *GAD1* for GABAergic interneuron; *AQP4* for astrocytes; *PDGFRA* for OPC; *CX3CR1* for microglia; *HBB* for blood cells and *FN1* for endothelial.

**Figure S3 Regional distribution of cells from ASD risk genes enriched subtypes.**

(A) Barplot displaying the regional composition of 4 ASD risk genes enriched subtypes. (B-D) Histogram showing the proportion of
(B) cells for subtype NPC-4 (B), Ex-1 (C), In-2 (D) and CR (E) from each brain region.

**Table S1. All gene with *de novo* mutations found in the Chinese ASD cohorts.**

**Table S2. TADA-Denovo analysis for genes with *de novo* mutations.**

**Table S3. List of chromosomal segments with *de novo* copy number variations which including genes presented in the SFARI gene list.**

**Table S4. Demographic information of ABIDE-I participants.**

**Table S5. Seed-based functional connectivity (left hemisphere).**

**Table S6. Seed-based functional connectivity (right hemisphere).**

**Table S7. List of abbreviation for brain regions.**

### Supplemental Materials and methods

#### Samples and ethics statement

We analyzed a sample set consisting of 369 ASD probands and 706 parents from 353 pedigrees recruited from Department of the Child and Adolescent Psychiatry, Shanghai Mental Health Center. Of the families 15 are multiplex that have two ASD children and 338 are trios. The fourth edition of the Diagnostic and Statistical Manual of Mental Disorders (DSM-IV) were used for ASD diagnoses made by trained psychiatrists. We obtained assent from the Institutional Review Board (IRB), Shanghai Mental Health Center of Shanghai Jiao Tong University (FWA number 00003065, IROG number 0002202). Dr. Yi-Feng Xu approved and signed our study with ethical review number 2016–4. Written informed consent was obtained from parents in consideration of the fact that all patients were minors. All participants were screened using the appropriate protocol approved by the IRB.

#### Whole-exome sequencing

Genomic DNA extracted from blood samples were sequenced at Shanghai Biotechnology Corporation (SBC) and WuXi NextCODE on Illumina HiSeq sequencers using the Agilent SureSelect Human All Exon V5 exome capture kit. Some samples were sequenced at Euler Genomics on Illumina HiSeq sequencers using the IDT xGen Exome Research Panel v1 exome capture kit. 150 bp paired-end sequencing reads were aligned to human genome build 38 (GRCh38/hg38) using the Burrows-Wheeler Aligner (BWA)(1), Picard tools MarkIlluminaAdapters, SamToFastq and MergeBamAlignment (http://broadinstitute.github.io/picard/) aggregated into a BAM file. Per-individual coverages of the target regions calculated by Qualimap 2 are shown in Figure S1A(2). Picard tools MarkDuplicates, SortSam and SetNmMdAndUqTags was used for marking duplicates, sorting by chromosome coordinates and adding essential tags. Single nucleotide variants (SNVs) and insertions / deletions (INDELs) were jointly called across all samples using the Genome Analysis Toolkit (GATK) HaplotypeCaller 4.1.4.1(3). Variant call accuracy was estimated using the GATK Variant Quality Score Recalibration (VQSR) approach and GATK CNN (Convolutional Neural Network) Variant Filter. The VCF file (format v4.2) was produced by the Broad sequencing and calling pipeline with GATK version 4.1.4.1.

We included variant calls with PASS flag in the downstream analyses. Variants (SNVs and INDELs) were annotated on the basis of the hg38 database using VEP(4). By following the definition of calculated variant consequences by VEP, we classified variants into those having HIGH, MODERATE, LOW and MODIFIER impacts.

#### Population stratification using genotyping data of common exonic SNPs

To define a set of common exonic SNPs, we first selected variants that are; 1) on the InfiniumExome-24v1-1_A1 genotyping array, 2) with MAF > 0.05 in East Asian (EAS) population of ExAC(5) annotated by VEP and 3) biallelic in EAS. After combining the information of these SNPs in our cohort (OWN) with the data of the same SNPs in African (AFR), American (AMR), East Asian (EAS), European (EUR) and South Asian (SAS) individuals in the 1000 Genomes Project(6), we performed further filtering and linkage disequilibrium (LD)-based pruning using PLINK v1.9(7) with the following options and parameters; --maf (minor allele frequency) 0.05, --mind (maximum per-person missing) 0.2, --geno (maximum per-SNP missing) 0.2, --hwe (Hardy-Weinberg disequilibrium p-value) 1×10^-10^ and --indep (SNP window size, number of SNPs to shift and variance inflation factor threshold) 50 5 2. By using the data of 1064 SNPs that passed the filters described above, we performed multidimensional scaling with PLINK.

#### Identification of DNMs

#### We filtered out variant calls when one or more variant alleles were observed in unaffected parents of our cohort (N of individuals = 326). By using the information of the remaining variant calls, we extracted candidates for DNMs using GATK PossibleDeNovo, TrioDenovo(8), DeNovoGear(9). Candidate DNMs called by these three tools at the same time were then stratified into SNVs and INDELs. We selected 98 DNM calls by prioritizing HIGH impact DNMs and MODERATE impact DNMs into consensus damaging missense. (CD-missense) DNMs were defined as the variants predicted to be damaging by at least two of the seven prediction algorithms: SIFT(10), PolyPhen-2 HumVar(11), PolyPhen-2 HumDiv(11), LRT(12), MutationTaster(13), Mutation Assessor(14) and PROVEAN(15) annotated by dbNSFP4.0a(16, 17).

#### CNV Detection

CNVs were called with GATK PreprocessIntervals, CollectReadCounts, AnnotateIntervals, FilterIntervals, DetermineGermlineContigPloidy, GermlineCNVCaller, IntervalListTools and PostprocessGermlineCNVCalls based on a cohort mode pipeline detecting germline copy number variants. All CNVs were annotated to GRCh38/hg38 by VEP and AnnotSV(18).

#### Real-time Quantitative PCR Validation

To confirm *de novo* CNVs detected by WES, quantitative PCR (qPCR) was performed using DNA from probands, their parents and controls. The comparative CT method (delta-delta CT method) was used for relative quantification, with data normalized against an endogenous control sequence (glyceraldehyde 3-phosphate dehydrogenase, GAPDH) with two normal copies. Genomic DNA was amplified using SYBR Green (Thermo Fischer Scientific) and qPCR was performed on a StepOnePlus™ Real-Time PCR System (Applied Biosystems). Data were analyzed using R version 3.6.3 and pcr package(19). The primers used for qPCR are *TBR1* Forward GGGATGACGAATCAGTCAGA, *TBR1* Reverse TGGCTGGACTGAGAGAGGAG, *RAI1* Forward TCTCCAGGCCAGAAAGAAAA, *RAI1* Reverse TGAATGCCTGGAATGAATGA, *SHANK3* Forward TGCCTCACGGAGTTTTCTCT, *SHANK3* Reverse ATGCGGGACTTTATGCAAAC, *MECP2* Forward CACGGAAGCTTAAGCAAAGG, *MECP2* Reverse TCAAGCACACCTGGTCTCAG, *GAPDH* Forward ATCAAGAAGGTGGTGAAGCA, *GAPDH* Reverse TGACAAAGTGGTCGTTGAGG.

#### Statistical Analyses

We statistically evaluated the observed number of dDNMs in each gene using TADA-Denovo(20). We included IMPACT HIGH and CD missense mutations in the TADA-Denovo analysis. Parameters for this analysis were determined by following the TADA manual. Per-gene mutation rates for LOF and CD missense DNMs were obtained from mirDNMR based on sequence context(21).

**Single cell RNA-seq data processing**

To infer the gene expression pattern of ASD risk genes in human brain, two publically available scRNA-seq dataset were downloaded(22, 23) and assembled. The assembled gene-by-cell count matrix was then imported into R package Seurat(24) for downstream analysis. Briefly, the assembled count matrix was first loaded into function CreateSeuratObject to create a Seurat object followed by a Log-normalization process using function NormalizeData. Variable genes were then identified with function FindVariableFeatures. Next, the principal component analysis was performed by function RunPCA. Unbiased clustering was done with function FindNeighbors and FindClusters followed by Uniform manifold approximation and projection (UMAP) dimension reduction analysis with function RunUMAP.

**Identification of differentially expressed genes**

Differentially expressed genes were computed with Seurat function FindAllMarkers by setting parameter only.pos =TRUE. Genes with adjusted P-values <0.05 were selected as differentially expressed genes.

**Enrichment analysis of ASD risk genes**

To explore the expression pattern of ASD risk genes, gene-set enrichment analysis was performed using R package AUCell(25) for the 55 High-impact genes and the 165 Moderate-impact genes on the assembled scRNA-seq dataset independently. Z-score of the gene-set enrichment score was then computed with R function scale and visualized by UMAP.

**Ranking of cells by enrichment score of ASD risk genes**

To study the expression pattern of ASD risk genes among different cell types and brain regions, we computed averaged gene-set enrichment score for each cell type and brain region and then ranked the cell types and brain regions in descending order. The averaged gene-set enrichment score and rankings were visualized with Heatmap.

**Proportion of highly expressed genes among subtypes and brain regions**

To get the proportion of highly expressed genes in each of the two gene-set (the 55 High-impact genes and the 165 Moderate-impact genes), for each gene-set, we counted the number of genes that have expression in at least 25% of the cells from each subtype or brain region. The proportion of highly expressed genes was then computed by dividing the number of highly expressed genes in each subtype or brain region by the total number of genes in each gene-set.

**Brain Imaging Data Acquisition**

Imaging data were obtained from Autism Brain Imaging Data Exchange (ABIDE-I, <http://fcon_1000.projects.nitrc.org/indi/abide/>)(26), a publicly available database released in 2012. 1112 subjects (539 ASDs, 573 age-matched healthy controls) from 16 international imaging sites underwent anatomical and resting-state functional MRI scans and a series of out-of-scanner phenotypic information was also collected. The scanning parameters of each site can be found online.

**Preprocessing of Imaging data**

All the preprocessed BOLD time series data and voxel-based morphometry (VBM) data used in this study was available on Preprocessed Connectomes Project (PCP; <http://preprocessed-connectomes-project.org/>). In this paper, resting-fMRI data and T1-weight MRI data from ABIDE were preprocessed by the Data Processing Assistant for Resting-State fMRI (DPARSF, <http://rfmri.org/dpabi>)(27) pipeline, one of the four preprocessing pipelines adopted by the ABIDE Preprocessed repository; further information can be found in the literature of Craddock et al.(28).

*Voxel-Based Morphometry*

For structural MRI, preprocessing including segment, spatial normalization, modulation and spatial smoothness. The raw T1w images of each subjects were segmented into gray matter (GM), white matter (WM), and cerebral-spinal fluid (CSF) tissue classes by calling the VBM8 toolbox (http://dbm.neuro.uni-jena.de/vbm/)(29) and transformed from individual native space into the Montreal Neurological Institute (MNI) space using the DARTEL normalization method(30). Then, normalized gray matter data were modulated to obtain the grey matter volume (GMV) measure. Finally, GMV data were smoothed with an 8 mm Full-width-Half-Maximum (FWHM) Gaussian kernel. Total brain volume (TBV = GM +WM) was calculated, which would be used as covariable in further analysis. According to the quality control information given in the phenotype file provided by the PCP, all scans rated as low quality (qc_anat_rater_1="fail" or qc_anat_rater_2="fail") were excluded for analysis.

We abstracted GMV of 6 regions of interest (ROI) for the statistical analysis: bilateral BST, PC and PRC. All ROIs were defined by the Desikan parcellation(31), and GMV of each ROI was obtained by taking average of all voxels located in the ROI.

*Seed-based Functional Connectivity*

For resting-state fMRI, preprocessing including removing the first 4 volumes, slice timing correction and head motion correction, regression of covariables, low-frequency filtering, spatial registration, spatial normalization and smoothness(27). We used a rigid body transformation to realign the time series of images for each subject. To reduce the respiratory and cardiac effects, signals from the WM and CSF are regressed out form the BOLD signals as covariables. Since the global signal is considered to reflect respiratory, cardiac and motion information which may led to spuriously correlation(32, 33), we performed the global signal regression to remove these potential confounding. Then, linear and quadratic trends were also regressed out, and a band-pass filtering (0.01–0.1 Hz) was performed. The transformation information acquired from T1w images normalization was used to perform spatial normalization for resting-state images. Finally, we performed a smoothness using a 6 mm FWHM kernel. All the subjects with low quality (qc_func_rater_1="fail" or qc_func_rater_2=="fail") or large head-motion (mean framewise displacement (FD) > 0.2 mm) were excluded.

Seed-based functional connectivity (FC) of the bilateral BST, PC and PRC was calculated. For each participant, we extracted averaged timing series within all ROIs and calculated their temporal coupling with all other brain voxels using Pearson correlation, resulting in 6 FC maps.

**Statistic**

For ROI-level GMV, we examined the differences between the ASD group and the health control group using a linear mixed model (LMM, using MATLAB built-in function *fitlme*), which has been shown to effectively control for site effects in previous large-scale neuroimaging studies. For the GMV of each ROI, we set the model as follows:

*GMV~Diagnosis+Sex+Age+TBV+(1|Site)*

Where the site effect was put into model as a random intercept, while diagnosis, age and sex were used as fixed effects. T-value and p-value of regression coefficient of *Diagnosis* were obtained from LMM to reflect the GMV difference between ASD and HC. For all statistical results, we applied a significant threshold of P < 0.05 (two-tails) and corrected for multiple comparisons using Family-Wise Error (FWE) method (number of comparisons=6).

For seed-based FC, we used a two-sample t-test to detect inter-group difference. Age, sex, site (coded as dummy variables), and head motion (mean FD) were regressed out as covariates before inter-group comparison. The t statistical maps were enhanced using the threshold-free cluster enhancement (TFCE)(34) with nonparametric permutation testing, which has been proven to be able to effectively increase reproducibility and control of false positive rate in voxel-level neuroimaging analysis(35, 36). The TFCE was performed based on the statistical module of the DPARSF toolbox. The number of permutations was set to 5000 and cluster-extent threshold of Z = 2.3. We applied a threshold of P < 0.05 (two-tails) and corrected for multiple comparisons (FWE) based on the results of a permutation test using the TFCE value.

**Additional Analysis**

To confirm that our results were not due to medication use, we excluded all subjects on medication and repeated our analysis. Besides, we repeated our analyses using full-scale intelligence quotient (FIQ) as a covariate to control the potential confounding effects from subject’s intelligence. The Intelligence in the Abide-I data set is mainly measured by Wechsler Instruments (Wechsler Adult Intelligence Scale (WAIS)(37) and Wechsler Intelligence Scale for Children (WISC)(38)), while there were also some other series scales used, refer to [http://fcon_1000.projects.nitrc.org/indi/abide/](http://fcon_1000.projects.nitrc.org/indi/abide/ABIDE_LEGEND_V1.02.pdf) for more information.
