## Supplemental Table 4 for "Identification of *de novo* mutations in the Chinese ASD cohort via whole-exome sequencing unveils brain regions implicated in autism"

Table S5. Demographic information of ABIDE-I participants.

| | | VBM | resting-state fMRI |
|---|---|---|---|
| ASD | N | 330 | 265 |
| | Sex (M) | 286 | 232 |
| | Age (Mean±SD) | 17.77±8.7 | 17.81±8.1 |
| | Medication | 43 | 58 |
| | FIQ | 105.95 | 107.74 |
| Healthy Control | N | 437 | 379 |
| | Sex (M) | 351 | 300 |
| | Age | 17.15±7.2 | 17.46±7.67 |
| | Medication (Other illness) | 2 | 1 |
| | FIQ | 110.96 | 111.41 |
