## Supplemental Table 5 for "Identification of *de novo* mutations in the Chinese ASD cohort via whole-exome sequencing unveils brain regions implicated in autism"

**Table S5. Seed-based functional connectivity (left hemisphere).**

The table lists brain regions that show significant FC differences between ASD and healthy control groups, where TFCE value represents the T value adjusted by non-threshold enhancement technique, and Corrected P value represents the P value adjusted by FWE correction. Cluster size indicates the size of the cluster. Only clusters significant in permutation test and bigger than 20 voxels (3*3*3*20=540mm^3^) were shown.

| left BST | | | | | | | |
| --- | --- | --- | --- | --- | --- | --- | --- |
| Area | MNI coordinates | | | T value | TFCE value | Corrected P value | Cluster size (mm3) |
|  | x | y | z |  |  |  |  |
| Cuneus/Precuneus | -6 | -78 | 30 | -4.79 | 609.549 | 0.0048 | 11151 |
| Superior Temporal/Transverse Temporal (R) | 60 | -24 | 12 | -5.50 | 567.863 | 0.008 | 2349 |
| Superior Temporal/Heschl (R) | 57 | -6 | 3 | -4.27 | 461.693 | 0.0206 | 297 |
| Precuneus (R) | 24 | -81 | 48 | -4.23 | 480.983 | 0.016 | 999 |
| left PC | | | | | | | |
| Area | MNI coordinates | | | T value | TFCE value | Corrected P value | Cluster size (mm3) |
|  | x | y | z |  |  |  |  |
| Fusiform (L) | -45 | -48 | -21 | -4.80 | 535.973 | 0.0108 | 2889 |
| Fusiform (R) | 45 | -51 | -24 | -4.48 | 484.363 | 0.017 | 1026 |
| Precuneus/Cuneus/Lingual (L) | -15 | -72 | 21 | -5.37 | 780.266 | 0.0006 | 26649 |
| Middle Temporal (L) | -48 | -48 | 9 | -4.64 | 502.022 | 0.0144 | 1647 |
| Insula (R) | 36 | -21 | 15 | -6.18 | 688.107 | 0.0022 | 7074 |
| Inferior Parietal (L) | -42 | -33 | 24 | -4.79 | 522.421 | 0.0126 | 2106 |
| Postcentral | 3 | -36 | 57 | -6.38 | 1359.3 | 0.0002 | 88614 |
| left PRC | | | | | | | |
| Area | MNI coordinates | | | T value | TFCE value | Corrected P value | Cluster size (mm3) |
|  | x | y | z |  |  |  |  |
| Precuneus (R) | 18 | -63 | 15 | -5.38 | 582.154 | 0.0072 | 10125 |
| Insula/Heschl (R) | 33 | -21 | 15 | -5.16 | 607.654 | 0.0058 | 5238 |
| Middle Temporal (L) | -63 | -48 | 12 | -4.33 | 477.922 | 0.0194 | 1647 |
| Middle Temporal (R) | 48 | -39 | 0 | -4.74 | 485.143 | 0.0188 | 459 |
| Inferior Parietal (L) | -39 | -39 | 21 | -4.11 | 476.775 | 0.0194 | 1134 |
| Precentral/Postcentral | 6 | -30 | 57 | -5.41 | 834.612 | 0.0004 | 60156 |
