## Supplementary figures and images for "Identification of *de novo* mutations in the Chinese ASD cohort via whole-exome sequencing unveils brain regions implicated in autism"

### Supplemental Table 7

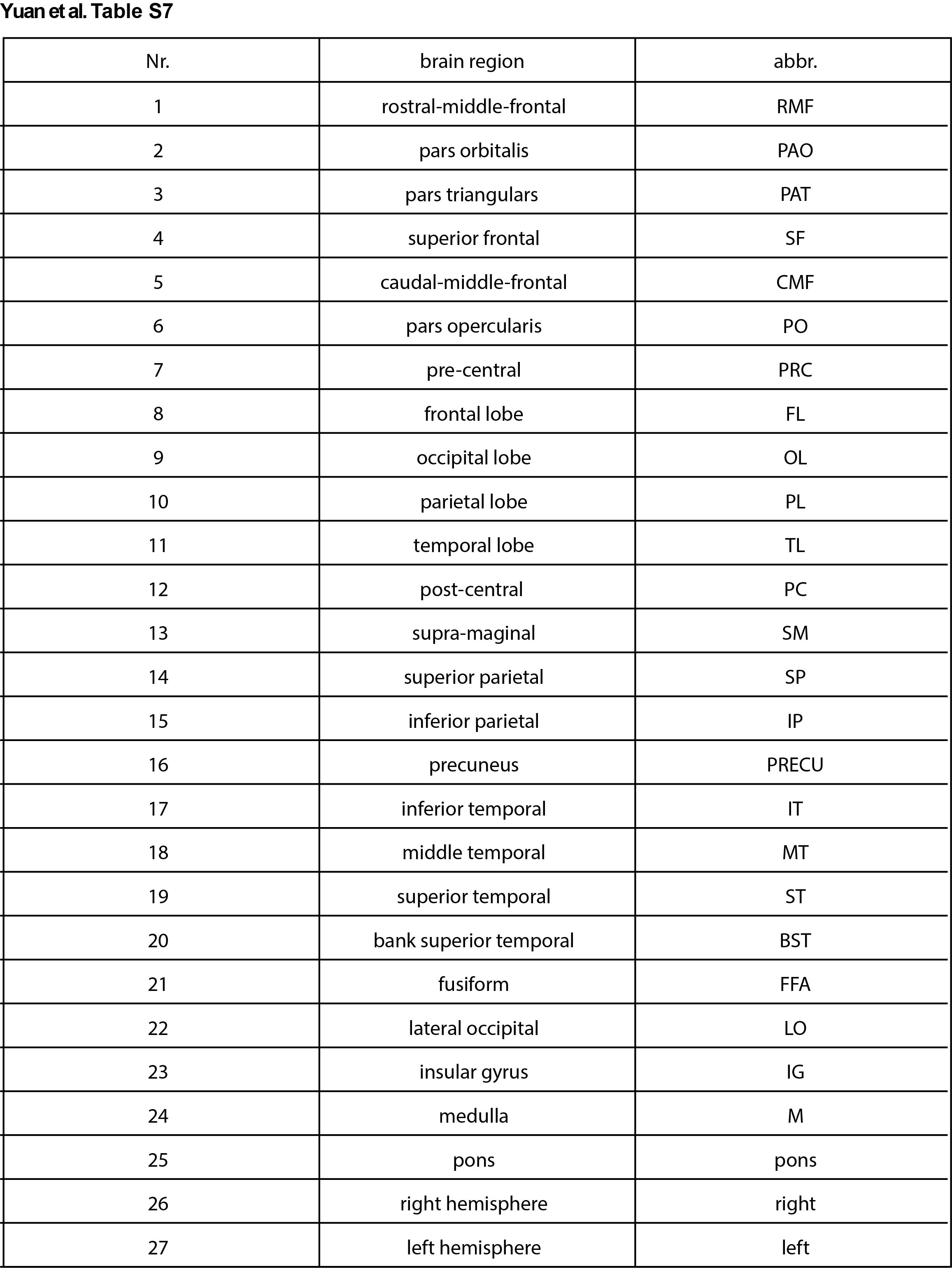
